# Children hospitalized for COVID-19 during the first winter of the pandemic in Buenos Aires, Argentina

**DOI:** 10.1101/2020.11.05.20225300

**Authors:** Silvina Raiden, Hector Cairoli, Javier Potasnik, Sandra Di Lalla, María José Chiolo, Fernando Torres, Paula Dominguez, Fernando Ferrero

## Abstract

**Background:** Although there are reports on COVID-19 in pediatrics, it is possible that the characteristics of each population, their health systems and how they faced the pandemic made the disease show distinctive features in different countries.

**Objective:** We aimed to describe the characteristics of patients hospitalized for COVID-19 in a tertiary pediatric hospital in the City of Buenos Aires, Argentina.

**Methods:** Descriptive study, including all patients hospitalized for COVID-19 in a tertiary pediatric hospital, from 04/26/2020 to 10/31/2020. Demographic, clinical and epidemiological characteristics of the patients are described.

**Results:** In the studied period 578 patients were hospitalized for COVID-19. The median age was 4.2 years and 83% had a history of close contact with a confirmed COVID-19 case. Regarding severity, 30.8% were asymptomatic, 60.4% mild, 7.4% moderate, and 1.4% severe. Among those with symptoms, the most frequent was fever, followed by sore throat and cough.

**Conclusion:** We reported 578 cases of children and adolescents hospitalized for COVID-19, most of them showed a mild or asymptomatic condition.

## INTRODUCTION

Despite the COVID-19 pandemic has caused hundreds of thousands of deaths worldwide, the information available to date shows that the disease is less severe in the pediatric population [1,2].

Also, there are reports that seem to show that the development of the pandemic may be influenced by the characteristics of each country, its health systems and how they faced the pandemic [3,4].

In addition to the characteristics of its own population and health system, Argentina faced the pandemic in a particular way, including the longest lockdown [5] and mandatory hospitalization of affected infants, regardless of its severity [6]. These characteristics could give a distinctive feature to children hospitalized for this disease.

We aimed to describe the characteristics of patients hospitalized for COVID-19 in a pediatric tertiary hospital in the City of Buenos Aires.

## METHODS

Retrospective study, including all patients hospitalized for COVID-19 at the tertiary pediatric hospital in Buenos Aires, Argentina,from April 26 to October 31, 2020. Besides patients admitted for COVID-19, all cases which require hospitalization for other reasons were also tested. The diagnosis was made by identification of SARS-CoV-2 in nasopharyngeal secretions by RT-PCR.

In all cases, sex, age, place of residence (including whether they lived in a poor neighborhood [7]), time of symptoms onset on admission, close contact with a confirmed COVID-19 case, presence of comorbidity and length of hospitalization was registered.

Severity was assessed as described by Dong [8]; patients presenting with SARS-CoV-2- related multisystemic inflammatory syndrome (MIS-C) were considered severe/critical.

Lab tests were also recorded (hemoglobin, differential WBC count, platelets, C-reactive protein, and erythrocyte sedimentation rate).

### Statistical analysis

Categorical variables are described by proportions with 95% confidence intervals (95%CI), and continuous variables by mean and standard deviation or median and interquartile range (IQR), according to distribution (Kolmogorov-Smirnov test). IBM SSPS Statistics 20.0 was used.

### Ethics

The study was approved by the Institution Ethics Committee.

## RESULTS

In the studied period, 578 children and adolescents were hospitalized for COVID-19. Some characteristics of 191 of these patients were mentioned in a preliminary report in the beginning of the pandemic [9]. Here we describe all patients hospitalized for COVID during the whole cold season (May-October), including laboratory data.

The number of hospitalizations per week ranged between 2 and 42 (Median= 30.5; IQR= 21.7-35.7) (Fig. 1).

**Figure 1.**
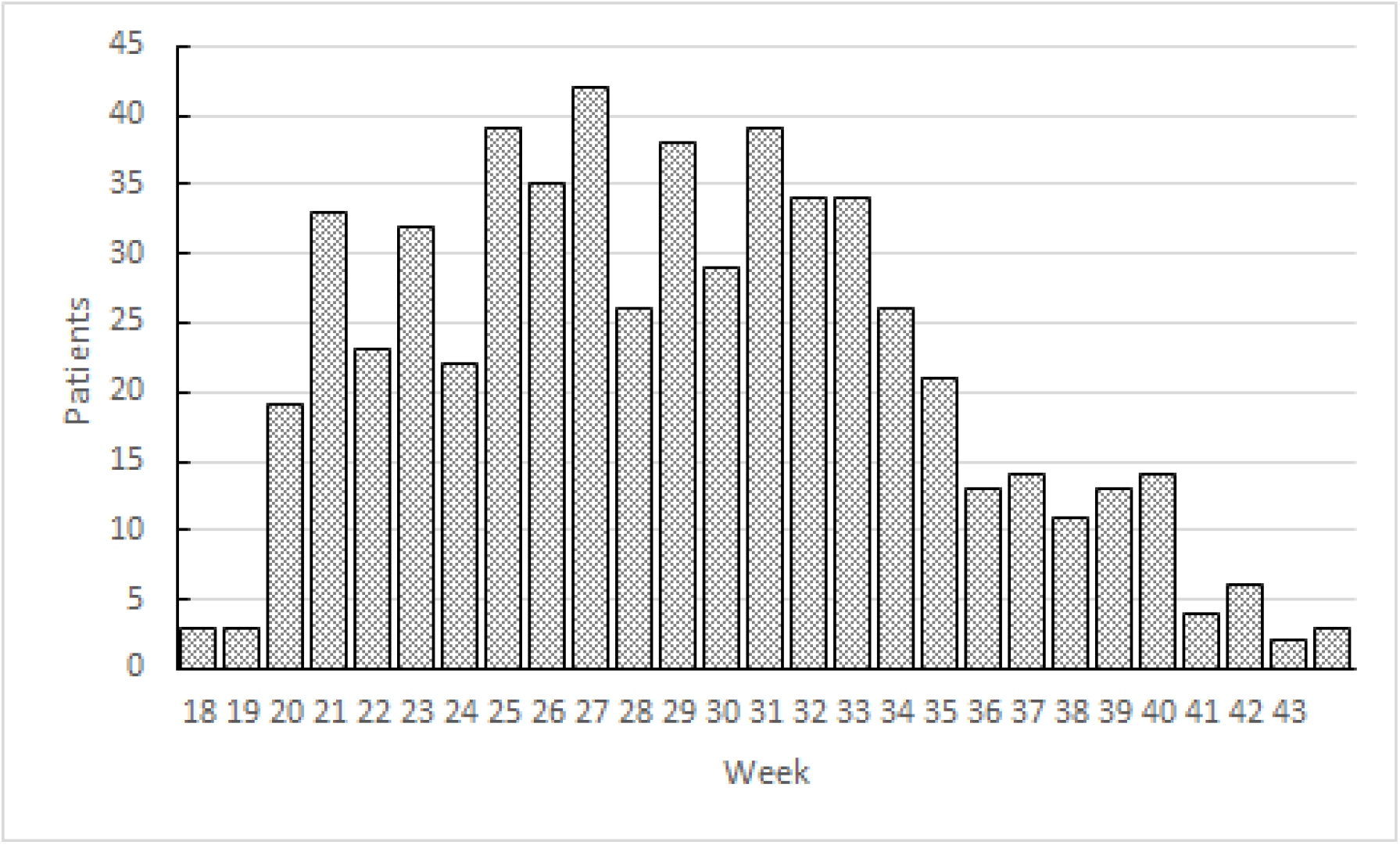
Pediatric hospitalizations for COVID-19, according to epidemiological week.

The median age was 4.2 (IQR: 0.7-11.2) years and 54.5% were male. Sixty-seven percent (67.3%) had their residence within the jurisdiction of the hospital (City of Buenos Aires) and, of them, 23.1% lived in a poor neighborhood.

Eighty-three percent (83%) had a history of close contact with a confirmed COVID-19 case. The time of symptoms onset before admission was 1 day (IQR: 1-3). Thirty-five percent (35.3%) had a previous or concomitant diagnosis of other disease, asthma being the most frequent (n = 49) (Table 1).

**Table 1.** Comorbidities in pediatric patients hospitalized for COVID-19 (n=204/578)

| **Disease** | **n** |
| --- | --- |
| Asthma | 49 |
| Surgical conditions (appendicitis, testicular torsion, hernias) | 12 |
| Seizures | 11 |
| Non-progressive chronic encephalopathy | 11 |
| Genetic disorders | 9 |
| Tumors | 8 |
| Tuberculosis | 8 |
| Other infections (otitis, celulitis, hepatitis) | 8 |
| Urinary tract infection | 7 |
| Obesity | 5 |
| Diabetes mellitus | 5 |
| Thrombocytopenic purpura | 5 |
| Chronic kidney disease | 5 |
| Chronic lung disease/Bronchopulmonary dysplasia | 4 |
| Congenital heart disease | 3 |
| Hemolytic-uremic syndrome | 2 |
| Hematologic disease (spherocytosis, hemophilia) | 2 |
| HIV | 2 |
| Other | 48 |

Disease severity was assessed and 30.8% were considered asymptomatic, 60.4% mild, 7.4% moderate, and 1.4% severe. From the six severe cases, only one required assisted ventilation, and two presented with SARS-CoV-2-related multisystemic inflammatory syndrome (MIS-C). Other six MIS-C cases were admitted to the hospital but SARS-CoV-2 testing was negative at that moment.

The most frequent initial symptom was fever, followed by sore throat and cough (Table 2). Regarding the lab tests, we found that 9.4% showed lymphopenia (lymphocyte differential count <20%) and 23.8% elevated C-reactive protein values (C-reactive protein > 10 mg/dl) (being 100% among severe cases and 10% among asymptomatics)(Table 3).

**Table 2.** Initial symptom of symptomatic patients hospitalized for COVID-19 (n=400)

| Symptom | n | % |
| --- | --- | --- |
| Fever | 207 | 51.7 |
| Sore throat | 49 | 12.2 |
| Cough | 40 | 10 |
| Rhinorrhea | 39 | 9.7 |
| Headache | 37 | 9.2 |
| Diarrhea/vomiting | 35 | 8.7 |
| Dyspnea | 17 | 4.2 |
| Abdominal pain | 15 | 3.7 |
| Loss of smell | 13 | 3.2 |
| Rash | 6 | 1.5 |
| Loss of taste | 3 | 0.7 |

(patients could have more than one symptom)

**Table 3.** Laboratory characteristics of children hospitalized for COVID-19.

|  | **Mean** | **SD** |
| --- | --- | --- |
| Hemoglobin (mg/dL) | 12.5 | 1.4 |
| White blood cells count (/mL) | 8,062 | 3,889 |
| Lymphocyte differential count (% ) | 49 | 20 |
| Neutrophil differential count (% ) | 39 | 20 |
| Platelet count (/mL) | 289,544 | 106,073 |
| Erythrocyte sedimentation rate (mm/1 h) | 13 | 12 |
| C-reactive protein (mg/dL) | 14 | 38 |

The length of stay had a median of 6 days (IQR: 2.2-9).

At the time of this report, only one patient with COVID 19 died, due to causes other than the disease, since he was in end-of-life care. All patients were discharged without complications (with phone follow-up), except those whose comorbidity prevented it.

## DISCUSSION

Our study strengthens the idea that COVID 19 in pediatrics is, in general, mild. It also supports some characteristics of the disease in Argentina that our early report has suggested [9].

Although it is possible that the age structure (Argentina has twice the number of children under 15 years of age than Italy) and the social behaviour of the different populations influence the differences observed [10] [11], the way in which local health authorities dealt with the pandemic have generated some particular features in our patients: Buenos Aires reported more pediatric cases than other countries, and we found a higher proportion of asymptomatic and mild among those hospitalized.

The City of Buenos Aires, adopted a “test and trace” policy. This program was carried out more strongly in poor neighborhoods, which have a younger population [12]. Mandatory hospitalization was also established for infants, still asymptomatic [6] [13]. Furthermore, those asymptomatic children and adolescents who did not have home conditions to comply with out-of-hospital isolation, were hospitalized for the isolation period [14].

In the period covered by this report (until 10/31/2020) the City of Buenos Aires reported 147,363 cases, of which 12.1% were children and adolescents [15], a substantially higher proportion than that initially reported in China (2.1 %) [16] and Italy [17], but closer to that lately reported in the USA (9%) [18].

We found a high proportion of asymptomatic patients (30.8%), higher than that reported by Götzinger in Europe in 582 children with SARS-CoV-2 infection (16%) [19], probably related to the above mentioned test and trace and institutional isolation policies adopted by the local health authorities. Despite this high number of asymptomatic children, the proportion of those with co-morbidity (35,3%) was similar to other series [20][21]. We only found 5.6% moderate and 1.2% severe cases. Dong et al. in China reported 5.9% severe and critical cases [8] and Tagarro in Spain reports that 9.7% of the cases in his series were severe [22]. Although a report from the United States showed that 32% of hospitalized pediatric patients with COVID-19 were admitted to a PICU, the limited average length of stay (2 days) and the limited proportion of subjects who required assisted ventilation (5.8%) suggests that the admission criteria in this series were broader [21].

Regarding the cases of MIS-C (COVID-19 associated Multisystem Inflammatory Syndrome in Children) that we report, they presented 4 to 6 weeks after reaching a significant number of cases in our city, a moment described as usual for the development of this complication [23]. Finally, we have found that 83% of our patients had close contact with infected people, supporting the idea that children are usually infected from adults [24].

This study has the limitation of presenting data from a single center, although it is probably the public institution that has hospitalized the most pediatric patients with COVID-19 in our country. Moreover, including data for a single center shows that, in a winter with no RSV related hospitalizations, the number of patients hospitalized for COVID-19 is roughly the same number we admit for RSV disease each year [25], whether this is related to non-pharmaceutical interventions used to control the pandemia or not [26].

## CONCLUSION

In this study we report 578 cases of children and adolescents hospitalized for COVID-19 in Argentina. Most presented their disease as mild or asymptomatic, supporting the idea that the management of pediatric COVID-19 patients represents more an organizational challenge rather than any one specific clinical task [27].

## Data Availability

The data that support the findings of this study are available from the corresponding author upon reasonable request.

## Notes

### Competing Interest Statement

The authors have declared no competing interest.

### Funding Statement

No external funding was received

### Author Declarations

Research Ethics Committee from the Hospital de Ninos Pedro de Elizalde has oversaw and approved this study

### Summary of Updates

Manuscript reviewed

## REFERENCES

1. Ludvigsson JF. Systematic review of COVID-19 in children shows milder cases and a better prognosis than adults. Acta Paediatr. 2020;109(6):1088-1095. doi:10.1111/apa.15270

2. Hoang A, Chorath K, Moreira A, et al. COVID-19 in 7780 pediatric patients: A systematic review. EClinicalMedicine. 2020;24:100433. Published 2020 Jun 26. doi:10.1016/j.eclinm.2020.100433

3. Smit AJ, Fitchett JM, Engelbrecht FA, Scholes RJ, Dzhivhuho G, Sweijd NA. Winter Is Coming: A Southern Hemisphere Perspective of the Environmental Drivers of SARS-CoV-2 and the Potential Seasonality of COVID-19. Int J Environ Res Public Health. 2020;17(16):5634. Published 2020 Aug 5. doi:10.3390/ijerph17165634

4. Tanne JH, Hayasaki E, Zastrow M, Pulla P, Smith P, Rada AG. Covid-19: how doctors and healthcare systems are tackling coronavirus worldwide. BMJ. 2020;368:m1090. Published 2020 Mar 18. doi:10.1136/bmj.m1090

5. Pieper O. Coronavirus: Argentina’s never-ending quarantine. Deutsche Welle. 2020 Aug 27. [cited 2020 Nov 01]. Available from: https://www.dw.com/en/coronavirus-argentinas-never-ending-quarantine/a-54721129.

6. Ministerio de Salud, Gobierno de la Ciudad Autónoma de Buenos Aires. Manejo frente a casos sospechosos y confirmados de coronavirus (COVID 19) en pediatría. 8 de Mayo de 2020. pp 6. [cited 2020 Nov 02]. Available from https://www.buenosaires.gob.ar/sites/gcaba/files/p.pediatria8.05.pdf.

7. Ministerio de Desarrollo Territorial y Hábitat. Registro Nacional de Barrios Populares. [cited 2020 Jun 25]. Available from: https://www.argentina.gob.ar/habitat/renabap.

8. Dong Y, Mo X, Hu Y, et al. Epidemiology of COVID-19 Among Children in China. Pediatrics. 2020;145(6):e20200702. doi:10.1542/peds.2020-0702

9. Cairoli H, Raiden S, Chiolo MJ, Di Lalla S, Ferrero F. Patients assisted at the Department of Medicine of a pediatric hospital at the beginning of the COVID-19 pandemic in Buenos Aires, Argentina. Arch Argent Pediatr 2020;118(6):418–426. Available from: https://www.sap.org.ar/uploads/archivos/general/files_cb_cairoli_eng_13-10pdf_1602265826.pdf

10. Natale F, Ghio D, Tarchi D, Goujon D, Conte A. COVID-19 Cases and Case Fatality Rate by age. [cited 2020 Nov 02] Available from: https://ec.europa.eu/knowledge4policy/sites/know4pol/files/jrc120420_covid_risk_and_age.pdf.

11. Dowd JB, Andriano L, Brazel DM, et al. Demographic science aids in understanding the spread and fatality rates of COVID-19. Proc Natl Acad Sci U S A. 2020;117(18):9696–9698. doi:10.1073/pnas.2004911117

12. Figar S, Pagotto V, Luna L, Salto J, Wagner Manslau M, Mistchenko A, Gamarnik A, Gomez Saldano AM, Quiros F. Community-level SARS-CoV-2 Seroprevalence Survey in urban slum dwellers of Buenos Aires City, Argentina: a participatory research. medRxiv 2020.07.14.20153858; [cited 2020 Nov 01]. Available from: https://www.medrxiv.org/content/10.1101/2020.07.14.20153858v2.

13. Ministerio de Salud, Gobierno de la Ciudad Autónoma de Buenos Aires. Manejo frente a casos sospechosos y confirmados de coronavirus (COVID 19). V.30. 7 de Junio de 2020. pp 6.

14. Ministerio de Salud, Gobierno de la Ciudad Autónoma de Buenos Aires. Manejo frente a casos sospechosos y confirmados de coronavirus (COVID 19). V.32. 19 de Junio de 2020. pp 16.

15. Ministerio de Salud, Gobierno de la Ciudad Autónoma de Buenos Aires. Actualización de los casos de coronavirus en la Ciudad. [cited 2020 Nov 01]. Available from: https://www.buenosaires.gob.ar/coronavirus/noticias/actualizacion-de-los-casos-de-coronavirus-en-la-ciudad-buenos-aires.

16. The Novel Coronavirus Pneumonia Emergency Response Epidemiology Team. The Epidemiological Characteristics of an Outbreak of 2019 Novel Coronavirus Diseases (COVID-19) — China, 2020[J]. China CDC Weekly, 2020; 2(8):113-122. [cited 2020 Nov 18]Available from: http://weekly.chinacdc.cn/en/article/id/e53946e2-c6c4-41e9-9a9b-fea8db1a8f51.

17. Livingston E, Bucher K. Coronavirus Disease 2019 (COVID-19) in Italy. JAMA.2020;323(14):1335. doi:10.1001/jama.2020.4344

18. CDC. CDC COVID Data Tracker: Demographic Trends of COVID-19 cases and deaths in the US reported to CDC. Atlanta, GA: US Department of Health and Human Services, CDC; 2020. [cited 2020 Nov 02]. Available from: https://www.cdc.gov/covid-data-tracker/index.html#demographics.

19. Götzinger F, Santiago-García B, Noguera-Julián A, et al. COVID-19 in children and adolescents in Europe: a multinational, multicentre cohort study. Lancet Child Adolesc Health. 2020;4(9):653–661. doi:10.1016/S2352-4642(20)30177-2

20. Rao S, Gavali V, Prabhu SS, Mathur R, Dabre LR, Prabhu SB, Bodhanwala M. Outcome of Children Admitted With SARSs-CoV-2 Infection: Experiences From a Pediatric Public Hospital. Indian Pediatr 2021; epub ahead of print. [cited 2021 Jan15] Available from: https://www.indianpediatrics.net/COVID29.03.2020/RP-00280.pdf.

21. Kim L, Whitaker M, O’Halloran A, et al. Hospitalization Rates and Characteristics of Children Aged <18 Years Hospitalized with Laboratory-Confirmed COVID-19 - COVID-NET, 14 States, March 1-July 25, 2020. MMWR Morb Mortal Wkly Rep. 2020;69(32):1081–1088. Published 2020 Aug 14. doi:10.15585/mmwr.mm6932e3

22. Tagarro A, Epalza C, Santos M, et al. Screening and Severity of Coronavirus Disease 2019 (COVID-19) in Children in Madrid, Spain [published online ahead of print, 2020 Apr 8]. JAMA Pediatr. 2020;e201346. doi:10.1001/jamapediatrics.2020.1346

23. Jiang L, Tang K, Levin M, et al. COVID-19 and multisystem inflammatory syndrome in children and adolescents. Lancet Infect Dis. 2020;20(11):e276–e288. doi:10.1016/S1473-3099(20)30651-4

24. Munro APS, Faust SN. Children are not COVID-19 super spreaders: time to go back to school. Arch Dis Child. 2020;105(7):618–619. doi:10.1136/archdischild-2020-319474

25. Ferrero F, Torres F, Abrutzky R, et al. Seasonality of respiratory syncytial virus in Buenos Aires. Relationship with global climate change. Arch Argent Pediatr. 2016;114(1):52–55. doi:10.5546/aap.2016.eng.52

26. Baker RE, Park SW, Yang W, Vecchi GA, Metcalf CJE, Grenfell BT. The impact of COVID-19 nonpharmaceutical interventions on the future dynamics of endemic infections [published online ahead of print, 2020 Nov 9]. Proc Natl Acad Sci U S A. 2020;202013182. doi:10.1073/pnas.2013182117

27. Parri N, Lenge M, Cantoni B, et al. COVID-19 in 17 Italian Pediatric Emergency Departments [published online ahead of print, 2020 Sep 23]. Pediatrics. 2020;e20201235. doi:10.1542/peds.2020-1235

